# Addressing spatial misalignment in population health research: a case study of US congressional district political metrics and county health data

**DOI:** 10.1101/2023.01.10.23284410

**Authors:** Rachel C. Nethery, Christian Testa, Loni P. Tabb, William P. Hanage, Jarvis T. Chen, Nancy Krieger

## Abstract

Areal spatial misalignment, which occurs when data on multiple variables are collected using mismatched boundary definitions, is a ubiquitous obstacle to data analysis in public health and social science research. As one example, the emerging sub-field studying the links between political context and health in the United States faces significant spatial misalignment-related challenges, as the congressional districts (CDs) over which political metrics are measured and administrative units, e.g., counties, for which health data are typically released, have a complex misalignment structure. Standard population-weighted data realignment procedures can induce measurement error and invalidate inference, which has prompted the development of fully model-based approaches for analyzing spatially misaligned data. One such approach, atom-based regression models (ABRM), holds particular promise but has scarcely been used in practice due to the lack of appropriate software or examples of implementation. ABRM use “atoms”, the areas created by intersecting all sets of units on which variables of interest are measured, as the units of analysis and build models for the atom-level data, treating the atom-level variables (generally unmeasured) as latent variables. In this paper, we demonstrate the feasibility and strengths of the ABRM in a case study of the association between political representatives’ voting behavior (CD-level) and COVID-19 mortality rates (county-level) in a post-vaccine period. The adjusted ABRM results suggest that more conservative voting record is associated with an increase in COVID-19 mortality rates, with estimated associations smaller in magnitude but consistent in direction with those of standard realignment methods. The results also indicate that ABRM may enable more robust confounding adjustment and more realistic uncertainty estimates, properly representing the uncertainties arising from all analytic procedures. We also implement the ABRM in modern optimized Bayesian computing programs and make our code publicly available, which may enable these methods to be more widely adopted.

## 1. Introduction

The COVID-19 pandemic, which simultaneously deepened political divisions in the United States (US) and shined a light on the role of the political system in population health, sparked increased scientific interest in the associations between political context and health outcomes^1– 5^. In contrast to assessments of individual political ideology and health, as measured by individual-level survey questions and/or by aggregated voting data (at the precinct or county level) on voter political lean, several members of our team recently published a novel study on the relationship between political representatives’ voting behavior and constituents’ health^5^. Specifically, this study analyzed associations between congressional representative voting behavior (plus other political metrics at the state level), and congressional district (CD) level COVID-19 outcomes reaggregated from county-level data. The analysis revealed that districts with representatives with more conservative voting patterns experienced increased COVID-19 mortality rates and stress on intensive care unit capacity, even after adjusting for sociodemographic and economic conditions and vaccination rates^5^.

Studies of this kind, which seek to link US congressional district-level political metric data with publicly available health data reported at standard census geographies (e.g., counties) must contend with the issue of areal spatial misalignment^6^. The same holds for other political units, such as state legislature districts, or city council or ward districts, which likewise can intersect with the boundaries of counties and census tracts. Areal spatial misalignment (hereafter simply *spatial misalignment* for brevity) occurs when areal data on multiple variables are collected using mismatched boundary definitions, and is a common obstacle to data harmonization and analysis across a range of fields. In political metric and health studies, political metric data may inherently arise at the CD level, e.g., representatives’ voting patterns, while the most granular health metric data publicly accessible are often aggregated to the county-level (as in Krieger et al^5^). Analyses seeking to investigate associations in CD and county-level features must confront a complex bi-directional spatial misalignment scenario, as counties can intersect CDs, be fully nested within CDs, or fully contain one or more CDs. Figure 1 displays an example of two types of spatial misalignment visible in the counties and CDs (from the 2010 redistricting cycle) in Vermont and New Hampshire. In Vermont, all counties are nested fully within the state’s single CD. In New Hampshire, there is non-nested misalignment in counties and CDs, with several counties intersecting both of the state’s two congressional districts.

**Figure 1.**
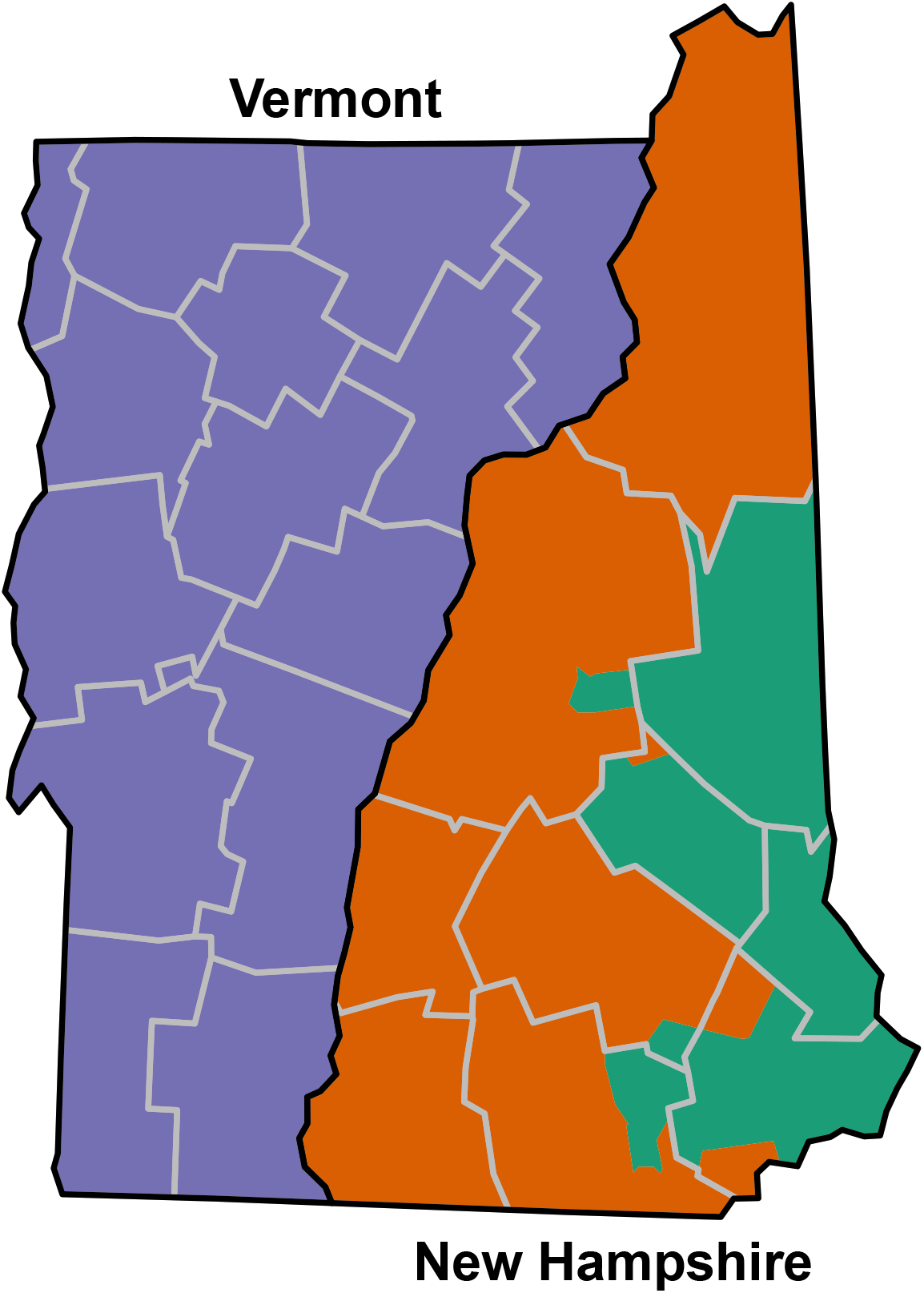
Example of two types of spatial misalignment in congressional district and county boundaries in New Hampshire and Vermont (using 2010 CD redistricting boundaries). The three US congressional districts in these states are shown in the three colors, county boundaries are grey, and state boundaries are black. In Vermont, all counties are nested fully within the state’s single congressional district. In New Hampshire, there is non-nested misalignment in counties and congressional districts, with several counties intersecting both the orange and the green congressional districts.

There is a large literature on the challenges and consequences of spatial misalignment^7–14^. In practice, spatial misalignment is nearly always dealt with during data processing, when each area-aggregated measure is reapportioned to a common target set of geographic units (“realigned”), and then analysis and inference proceeds as usual using the realigned data. Dasymetric methods represent one approach to realignment, where a strong and often unrealistic assumption of no within-area variability is imposed for each measure, and values are reapportioned to the target units via population-weighting. Dasymetric methods have previously been used to realign data in the context of political metrics and health research^5^. This type of apportionment-based realignment is deterministic and can introduce measurement error and confounding that bias effect estimates of interest^15,16^. Additionally, because the uncertainty introduced by the realignment procedures is typically not accounted for in downstream modeling, analytic results are likely to suffer from under-estimated uncertainties and invalid inference.

Fully model-based methods for addressing spatial misalignment have also been proposed in the statistical literature^6,9,11,13,17–19^. Many such methods approach the misalignment problem by assuming that any measure used in the analyses arises from a smooth underlying spatial risk surface^6,9,14,19^. Then, Gaussian process models are employed to create smooth predicted risk surfaces for each measure, and associations are assessed by extracting and analyzing predictions at a common set of points from each measure’s risk surface. In the context of political metrics and health research, because the scientific hypotheses contend that political representation and voting patterns are associated with health/health-relevant exposures^20^, and political representation may change sharply at congressional district boundaries, the notion of a smooth underlying spatial surface is at odds with the hypotheses.

These considerations motivate our focus on a small literature proposing atom-based regression models (ABRM) for analyzing spatially misaligned data^17,21^, which do not rely on the assumption of a smooth spatial risk surface. ABRM use as the units of analysis the areas created by intersecting all sets of areal units on which variables of interest are measured (“atoms”).

Because, in the most general case, variables of interest are unobserved at the atom level, this approach treats the atom-level unmeasured values of covariates or outcomes as latent variables, and builds models allowing them to be sampled, conditional on observed values across some unions of the atoms. ABRM provide a fully model-based approach for analyzing misaligned data without any assumption of smoothness or need to spatially align all variables prior to analysis, and the resulting inferential quantities properly represent the uncertainties arising from all model components, in contrast to deterministic reapportionment approaches. Thus, they offer a promising modeling framework for studying associations between political metrics and health.

Although they were introduced in the statistical literature more than two decades ago, ABRM have rarely, if ever, been employed in scientific practice. This is likely due to the difficulty of implementing these complex Bayesian models from scratch and the lack of “off-the-shelf” software or examples to guide implementation, as well as their sizeable computational demands. In this paper, we implement an ABRM in Nimble^22,23^, a modern, optimized Bayesian sampling software accessible through R^24^, and demonstrate its application to a case study of political metrics and health. In particular, we conduct an analysis that builds on the study by Krieger et al^5^ to assess the association between congressional representatives’ voting patterns (CD level) and COVID-19 mortality (county level), during a period of vaccine availability, using ABRM. We compare the ABRM results to those obtained using standard realignment approaches with various selections for the target units. This work can serve as a guide to others seeking to use model-based approaches to analyze spatially misaligned areal data, in the context of political metric and health studies, and beyond.

## 2 Methods

### 2.1 Spatial units of analysis

Our study is based on data for the entire continental US, to sidestep issues with spatial modeling that arise when including Alaska and Hawaii. The most spatially granular level at which the COVID-19 outcomes of interest (described in detail in Section 2.2) are released is the county level. However, our primary covariate of interest (Section 2.3) is inherently defined at the CD level, using CD boundaries from the 2010 redistricting cycle. We thus wish to assess associations between variables collected at the county and CD levels, which creates a two-level, bi-directional spatial misalignment structure (Figure 2). In the time period under study, while most counties--87% of counties in the continental US--were fully contained within a single CD, others were split by CD boundaries, and in a few rare cases largely occurring in densely populated counties, multiple CDs were fully contained within a single county. Moreover, while only 13% of counties were intersected by more than one CD, these counties contained 60% of the US population.

**Figure 2.**
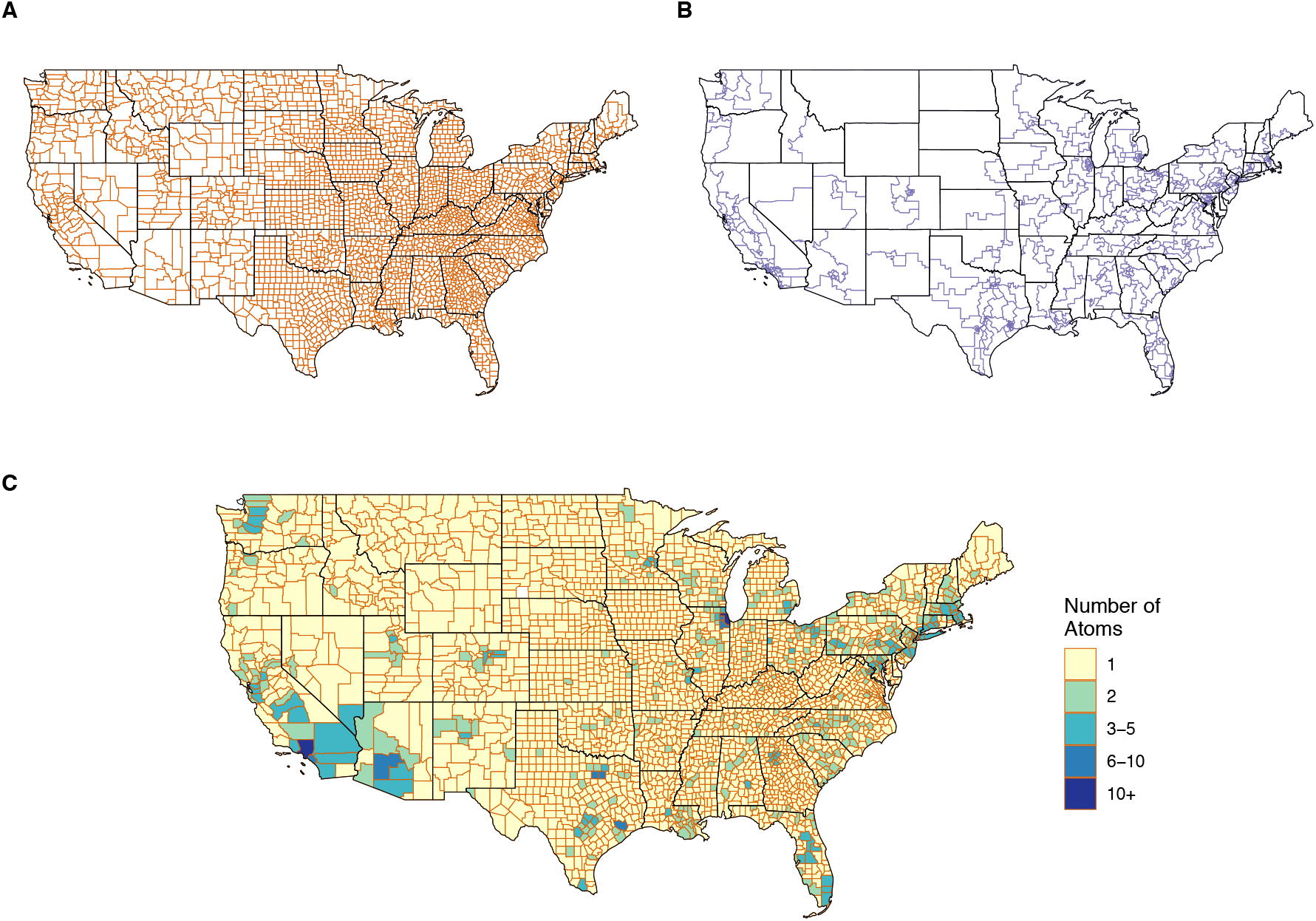
Continental US county boundaries (A), 2010 congressional district boundaries (B), and the number of atoms into which each county is split when the county and congressional district maps are intersected (C). The analytic dataset contains 3,104 counties, 432 congressional districts, and 3,728 atoms.

For this study, atoms are created by intersecting county and CD boundaries (Figure 2). Thus, each atom overlaps a single county and a single CD. Atoms containing zero population are omitted from the analyses (1.7% of atoms). Our analytic dataset contains 3,104 counties, 432 CDs, and 3,728 atoms (Table 1). Note that our exclusion of Alaska (1 CD) and Hawaii (2 CDs) accounts for our use of 432 out of the total 435 US CDs. 72% of the atoms are county-equivalent atoms, i.e., atoms exactly equivalent to counties, a result of the fact that most counties are fully contained within one CD. The remaining atoms are sub-county atoms.

**Table 1.** N and mean (standard deviation) for variables collected at the county level, congressional district (CD) level, and atom level, as available, for the continental US (2016-2022)*.

|  | County-level | CD-level | Atom-level |  |  |
| --- | --- | --- | --- | --- | --- |
|  |  |  | Overall | County-equivalent atoms | Sub-county atoms |
| N | 3104 | 432 | 3728 | 2686 | 1042 |
| Population size (in thousands) | 105.84 (337.14) | 760.37 (60.97) | 87.54 (152.73) | 48.81 (83.39) | 187.38 (227.45) |
| COVID-19 deaths<sup>†</sup> | 121.55 (306.55) |  |  | 70.96 (103.14) |  |
| DW-NOMINATE |  | 0.05 (0.46) | 0.32 (0.41) | 0.42 (0.33) | 0.07 (0.46) |
| Percent poverty<sup>‡</sup> | 13.90 (5.96) | 11.90 (4.05) |  | 14.20 (6.05) |  |
| Population density | 10.69 (72.08) | 96.73 (279.93) | 13.85 (56.96) | 3.09 (14.70) | 41.58 (99.95) |
| Percent Black | 8.80 (14.16) | 12.36 (13.3) | 9.27 (14.59) | 8.23 (14.08) | 11.96 (15.53) |
| Percent Hispanic | 9.83 (13.75) | 18.64 (17.81) | 11.08 (14.98) | 9.17 (13.52) | 16.00 (17.27) |
| Percent Asian | 1.36 (2.60) | 5.94 (7.30) | 2.06 (4.41) | 0.95 (1.56) | 4.91 (7.22) |
| Percent AIAN | 1.82 (6.58) | 1.12 (2.03) | 1.78 (6.84) | 1.92 (6.84) | 1.43 (6.82) |

\*Population size and racialized group composition data were obtained from the 2020 US Census; COVID-19 deaths and the DW-NOMINATE data are for the time period April 1, 2021-March 31, 2022; poverty was obtained from the 2016-2020 American Community Survey 5-year estimates.
<sup>†</sup>COVID-19 deaths are observed at the atom level for county-equivalent atoms only.
<sup>‡</sup>Percent poverty is observed at the atom level for county-equivalent atoms only and was imputed based on model (3) for sub-county atoms.

All data used in our analyses are collected at either the county level, CD level, or the census block level. Census blocks are the only census geographies that are guaranteed by design to be fully nested within both counties and CDs, so data obtained at the census block level can be directly aggregated to the atom level.

### 2.2 Outcomes

Our outcome of interest is COVID-19 mortality during April 2021-March 2022, a time frame during which the COVID-19 vaccine was publicly available in the US. Counties are the smallest units for which COVID-19 mortality data are made publicly available nationwide. As in Krieger et al^5^, we collect county-level COVID-19 death counts for April 2021-March 2022 from the CDC Wonder Database (Provisional Mortality Statistics). Death counts <10 are suppressed in this data source, thus we impute values for counties with suppressed counts as described in Krieger et al^5^.

Because our interest lies in COVID-19 age-adjusted death *rates* rather than death *counts*, we also obtain population count data to serve as denominators. Because our model is built at the atom level, we prefer atom-level data where available. Thus, we compute total population counts by aggregating census block-level counts from the 2020 Decennial Census redistricting files to the atom level. At the time of writing, age distribution data from the 2020 census have not yet been released, so we obtain age distribution information at the county level from the 2016-2020 5-year American Community Survey (ACS) data. We assume that age distributions are constant across atoms within each county and use these age distributions, alongside atom-level total population counts, to create atom-level expected COVID-19 mortality counts. These serve as the denominators in our models of age-standardized rates.

### 2.3 Predictors

Our primary covariate of interest is the DW-NOMINATE dimension score 1^25^, which is a measure of the political ideology of a CD’s congressional representatives calculated based on votes cast April 2021-March 2022. DW-NOMINATE can take values between -1 and 1, with lower values indicating more liberal voting patterns and higher values indicating more conservative patterns. This measure is defined and created at the CD level. Importantly, we note that it is not an aggregate measure--each individual in the CD is “exposed” to the voting patterns of the CD’s representatives—and is therefore constant within a CD. Thus, for the purposes of our atom-based modeling approach, we can assume that each atom should inherit the DW-NOMINATE score of the CD containing it.

As in Krieger et al^5^, the aim of this case study is to assess associations, not to make causal inference. However, we wish to adjust for several well-established community-level risk factors for COVID-19, and in this study use several social and economic variables also included in the Krieger et al study^5^. In particular, community impoverishment is an important adjustment variable in our analyses, which we measure using the percent of residents in poverty extracted from the 2016-2020 5-year ACS at the county level. Note that this variable can in fact be obtained down to the census block group level, but because census block groups are not nested within our other geographies (CDs and atoms), collecting data at that level would introduce another level of spatial misalignment, so we use the county-level measure instead. Poverty is an area-aggregate measure and so, unlike DW-NOMINATE, cannot be assumed to be constant across atoms within a county. We also adjust for areal racialized group composition measures (percent Black, percent Hispanic, percent Asian, and percent American Indian/Alaska Native [AIAN]) and population density in our models, which are collected at the census block level from the 2020 Decennial Census redistricting files and aggregated to atoms.

### 2.4 Atom-based regression model

Following the notation in Trevisani and Gelfand^21^, we let *i* = 1, …, *N* index counties and *k* = 1, …, *K* index atoms created by intersecting counties and CDs. We denote the observed county-level COVID-19 mortality count outcomes by *Y*_*i*_, the county-level poverty counts as *X*_*i*_, and the vector of the remaining, atom-level covariates (DW-NOMINATE, racial/ethnic composition, and population density) by *C*_*k*_. Let *P*_*k*_ represent the total population size of atom *k* and *E*_*k*_ represented the expected number of COVID-19 deaths for atom *k*. For the features observed at the county level, *Y*_*i*_ and *X*_*i*_, we also conceive of possibly latent atom-level values of these features, denoted by *Y*_*k*_ and *X*_*k*_. In the most general ABRM setting, atom level features can be entirely unobserved/latent. However, in our setting, the majority of atoms are county-equivalent atoms, and because *Y* and *X* are observed at the county-level, *Y*_*k*_ and *X*_*k*_ are observed for these county-equivalent atoms. For sub-county atoms, *Y*_*k*_ and *X*_*k*_ are latent.

We then postulate the following models for these atom-level variables

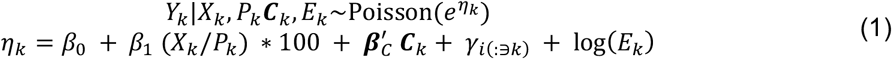

and

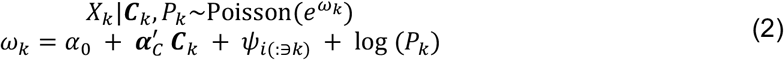

Here *γ*_*i*_ and *ψ*_*i*_ are county-level spatial random effects, and each atom inherits the random effect value of its corresponding county. Because here *Y*_*k*_ and *X*_*k*_ are latent for sub-county atoms-- and in more general ABRMs, atom-level features may be entirely latent-- we leverage the relationship between the atom-level variables and the observed data to learn about the parameters of models (1) and (2). Note that the observed county-level COVID-19 mortality counts, *Y*_*i*_, and poverty counts, *X*_*i*_, are sums of the counts across the atoms within each county. The atom-level measures are assumed to be Poisson distributed, and sums of Poisson random variables are also Poisson distributed. Thus, the model specifications at the atom level induce the following distributions for the fully-observed county-level data:

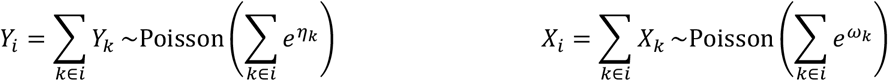

Estimation of the ABRM proceeds within a Bayesian framework. A joint likelihood is formulated based on these distributions, and prior distributions are specified for the parameters and random effects. Here, we specified intrinsic conditionally auto-regressive (ICAR) spatial priors for the random effects in each model, Normal(0,1) priors for the ***β*** and ***α*** parameters, and Gamma(0.001,0.001) priors for the precision hyperparameters in the ICAR priors.

The primary purpose of the model for *Y* is to estimate the coefficients =*β*_1_ and ***β***_***C***_, while the purpose of the model for *X* is to obtain predictions of the latent *X*_*k*_ values in sub-county atoms for downstream use in the model for *Y* (note that we can estimate the ***α*** coefficients from the model for *X*, but these are usually not of primary interest). While we could predict the latent *X*_*k*_ values directly from model (2), doing so without constraints will generally result in 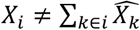, i.e., the sum of predicted atom-level values within a county is not equal to the observed county-level totals. To avert this issue and better utilize all information in the observed data, we obtain predictions for sub-county atoms conditional on the corresponding county totals, leveraging the fact that Poisson random variables conditional on their sum follow a Multinomial distribution. For sub-county atoms *m* = 1, …, *M*_*j*_ nested within county *j*, the latent *X*_*m*_ values can be sampled conditional on their corresponding county totals, *X*_*j*_, as

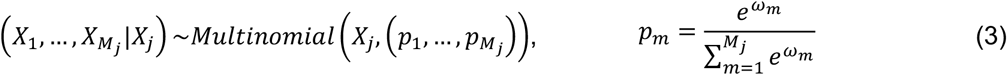

Note that the latent *Y*_*k*_ values for sub-county atoms can be sampled analogously if desired.

All model components are fit jointly via Markov Chain Monte Carlo (MCMC) sampling, enabling uncertainties to be fully propagated through all components. Code to implement the model was written in Nimble^22^ and executed through R. We report rate ratio estimates as exponentiated posterior means for the coefficient parameters and corresponding 95% credible intervals (CIs) from the models.

### 2.5 Analyses using standard methods

To compare the ABRM results to those that would be obtained using standard dasymetric realignment approaches (as described in Section 1), we conduct two additional analyses: a CD-level analysis and a county-level analysis. First, following Krieger et al^5^, we conduct a CD-level analysis in which we (1) use a dasymetric approach to realign county COVID-19 mortality rates to the CD level; (2) collect all adjustment variables at the CD level from the 2016-2020 ACS and 2020 census redistricting files; and (3) fit a Poisson regression model to the CD-level measures. Note that, while this analysis is similar to that of Krieger et al^5^, we use a smaller set of adjustment variables and more updated data sources that were not available at the time of their analyses (i.e., 2020 decennial census data). For the county-level analysis, we (1) assign each county the DW-NOMINATE score of the CD in which its centroid lies; (2) collect all adjustment variables at the county level from the 2016-2020 ACS and 2020 census redistricting files; (3) fit a Poisson regression model to the county-level measures. From each of these analyses, we report rate ratios and 95% confidence intervals.

## 3 Results

The final analytic dataset contains 3,104 counties in the continental US, 432 congressional districts, and 3,728 atoms (Figure 2, Table 1). There are 2,686 county-equivalent atoms and 1,042 sub-county atoms. Sub-county atoms tend to be concentrated in densely populated counties, which can contain multiple CDs. Illustrating this, county-equivalent atoms have an average population size of 48,810, while sub-county atoms (atoms falling in counties containing 2+ atoms) have an average population size of 187,378 (Table 1).

The COVID-19 mortality rate ratio (MRR) estimates and 95% CIs from the ABRM, CD-level model, and county-level model are shown in Figure 3, where all models use age-standardized outcomes and are adjusted for the additional covariates described in Section 2.3. In each of the models, all covariates besides poverty were standardized prior to modeling, so that 100*(MRR-1) can be interpreted in the percent change in COVID-19 mortality rate corresponding to a 1-standard deviation increase in the covariate. Poverty, which required special considerations in the ABRM due to its misalignment and cannot be easily standardized, was left on the percent scale in all models. With the exception of the percent Black variable, the MRR estimates from the three models agree in terms of direction, but the magnitude of the estimated associations from the three models often differs dramatically. Moreover, the uncertainties from the ABRM are consistently much larger than those from the CD- and county-level models. This is likely a result of the ABRM’s more comprehensive characterization of model-based uncertainty compared to approaches that rely on deterministic reapportionment, as well as possibly the smaller population sizes of atoms (smaller areas tend to have more variable disease/mortality rates).

**Figure 3.**
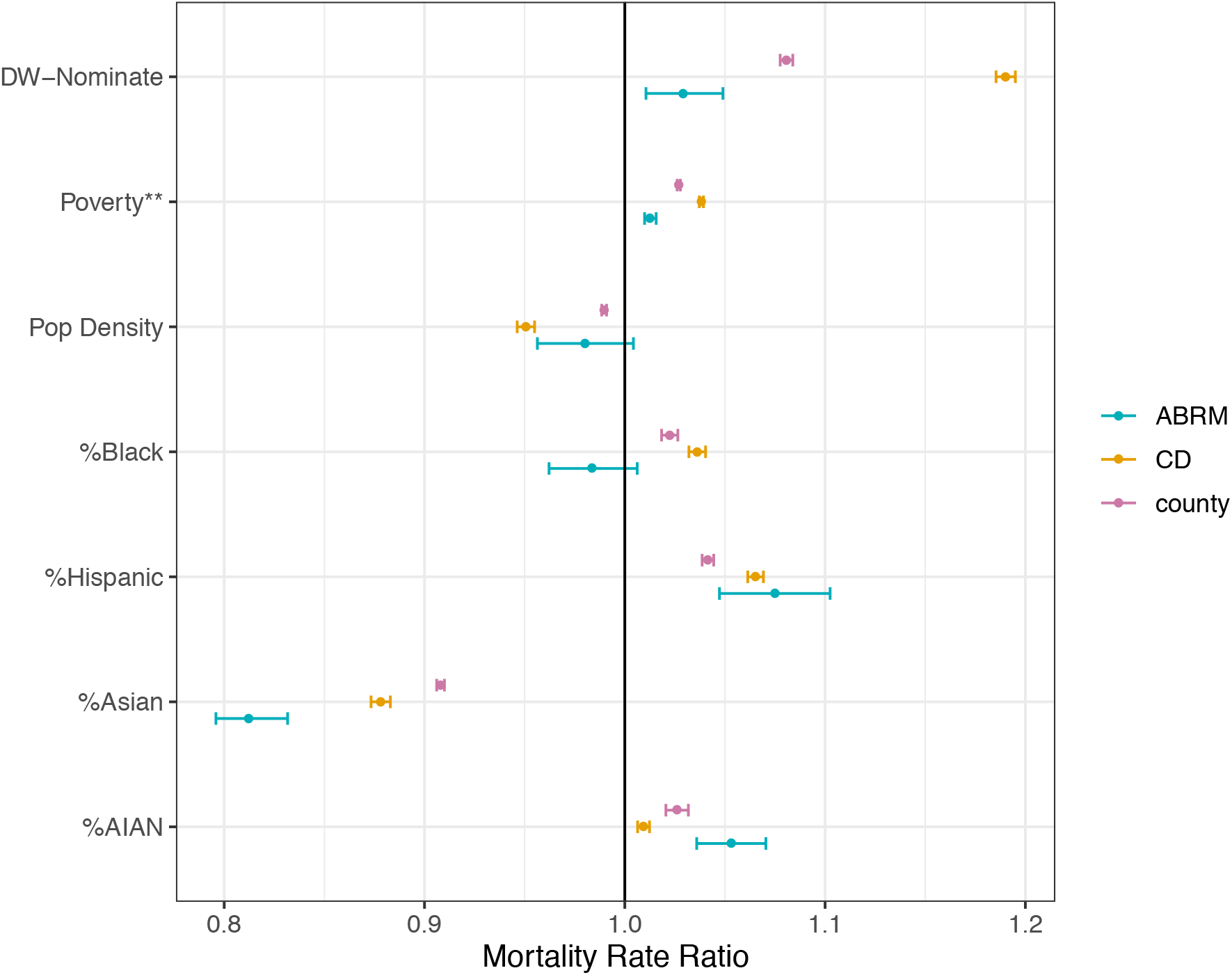
COVID-19 mortality rate ratio (MRR) estimates and 95% credible intervals from the atom-based regression model (ABRM), the congressional district-level model (CD) and the county-level model (county). Each model is fit to age-standardized COVID-19 mortality rate data and co-adjusted for all variables simultaneously. All measures besides poverty are standardized so that the MRR corresponds to a one-standard deviation increase in the coefficients. Poverty is on the percent scale and not standardized.

The ABRM found that a one standard deviation increase in DW-NOMINATE was associated with a 2.9% (1.1%, 4.9%) increase in COVID-19 mortality rates during April 2021-March 2022, indicating that an increase in conservative voting patterns by the area’s congressional representatives is associated with increased COVID-19 mortality rates, after conditioning on poverty, population density, and areal racialized-group composition. The MRR estimates from CD- and county-level models were much larger in magnitude. The CD-level model found a 19.0% (18.5%, 19.5%) increase in COVID-19 mortality rates and the county-level model found an 8.1% (7.8%, 8.4%) increase in COVID-19 mortality rates for every standard deviation increase in DW-NOMINATE.

Note that the trend in estimated DW-NOMINATE MRRs from the CD-level model, county-level model, and ABRM aligns with the general trend in population size of the units of analysis (on average, CDs > counties > atoms). The CIs for the DW-NOMINATE MRRs are widest for the ABRM, for reasons noted above, and narrowest for the county-level model. The wider CIs for the CD-level model compared to the county-level model are likely due to the much smaller number of CDs (N=432) relative to counties (N=3,104).

In all models, areas with increased percent poverty, percent Hispanic residents, and percent AIAN residents were found to have substantially higher COVID-19 mortality rates, while areas with increased percent Asian residents had lower rates (Figure 3). CD- and county-level models estimated positive associations between percent Black and COVID-19 mortality rates, while the ABRM estimated an inverse association, but with 95% CI containing the null value. For completeness, we also report the rate ratio estimates from the poverty model, i.e., 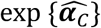 from model (2), and corresponding 95% CIs in Figure S1 of the SI, although they are not of substantive interest here.

To provide further insight into the performance of the ABRM, we mapped the model-estimated atom-level standardized COVID-19 mortality rates (SMRs) for Los Angeles County, California (LA County) in Figure 4 (left panel). LA County serves as an informative example of the model’s performance, as thanks to its large population (>10 million), it is split across 18 CDs and therefore contains 18 sub-county atoms. Moreover, the LA County Department of Public Health has released sub-county health district-level age-adjusted COVID-19 mortality rate maps (Figure 4, right panel)^26^, which although representing data for the entire pandemic through January 8, 2023 (with only this cumulative data available from the data dashboard^26^, as opposed to the more limited post-vaccine period investigated here), enables general comparisons of spatial patterns with our sub-county COVID-19 mortality rate estimates. While the absolute rate values in the two maps should not be compared, we can see that the ABRM estimates display similar spatial patterns to the observed rates, indicating that the ABRM is capturing key trends in COVID-19 mortality risk. Given dynamic temporal trends in both COVID-19 mortality rates and social and spatial inequities in these rates, for future analyses we are investigating feasibility of obtaining the LA sub-county health district-level age-adjusted COVID-19 mortality rates for solely the April 2021-March 2022 period, to match temporally to our vaccine-era analyses.

**Figure 4.**
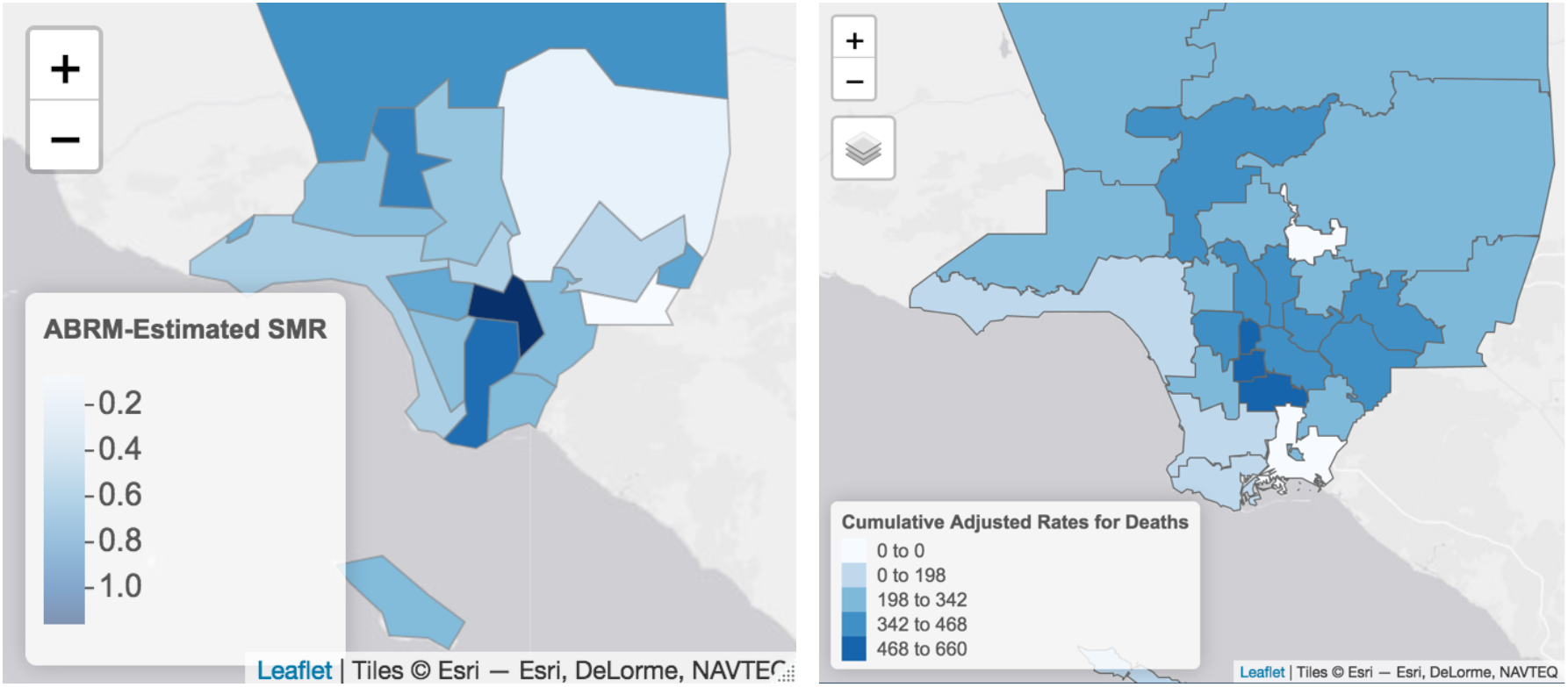
Left panel: Map of ABRM-estimated age-standardized COVID-19 mortality rates (SMRs) for April 2021-March 2022 across atoms in Los Angeles County, California. Right panel: age-adjusted observed COVID-19 death rates from the start of the pandemic through January 8, 2023 by health district from the LA County Public Health Department COVID-19 Surveillance Dashboard^26^.

## 4 Discussion

In this paper, we demonstrated the feasibility and the benefits of employing ABRM to assess associations between spatially misaligned variables using a case study of COVID-19 mortality rates and political representatives’ voting records in the post-vaccine period. As in Krieger et al^5^, who studied the same question using standard reapportionment techniques to handle the spatial misalignment of county-level and CD-level data, we found that a more conservative voting record of congressional representatives was associated with higher age-standardized COVID-19 mortality rates in a post-vaccine era, after adjusting for community-level poverty, racialized group composition, and population density. However, when compared to the results obtained with approaches that realign all data to the county or CD levels, the ABRM yields associations much smaller in magnitude with larger associated uncertainties.

By design, the ABRM learns about associations between variables at spatial units smaller than, or equivalent to, those on which the variables were observed, and this higher resolution analysis may enable improved confounding adjustment compared to the more aggregated analyses typically conducted when all data are a priori realigned to the spatial units on which some of the variables were observed (here, counties or CDs). In our case study, we find evidence of this phenomenon, as the magnitude of estimated association between representatives’ voting record and COVID-19 mortality moved towards the null with smaller spatial units of analysis (CD-level estimates > county-level estimates > ABRM estimates), which may well be due to more robust confounding adjustment. Moreover, the ABRM is able to fully account for uncertainties arising due to misalignment, as indicated by the larger CIs for the ABRM estimates relative to those from a priori realignment approaches, which ignore uncertainty arising from the realignment procedures and therefore likely under-estimate uncertainties and provide unreliable inference.

Given today’s vast amount of administrative data collected over incompatible geographies, the need for accessible methods that address spatial misalignment in a statistically-principled manner is greater than ever. ABRM are a powerful tool for assessing associations in misaligned data, but are often overlooked in practical applications, likely due to the absence of software or code examples to assist with implementation. Our work and publicly available code on Github (https://github.com/rachelnethery/atom_model) provide a prototype for implementation of ABRM leveraging modern optimized software for MCMC sampling, which can serve as a template to enable these methods to be more widely adopted in scientific research.

## Data Availability

All data used in the present study are publicly available and can be obtained from CDC Wonder (https://wonder.cdc.gov/mcd-icd10-provisional.html) and the US Census Bureau (https://data.census.gov/).

https://github.com/rachelnethery/atom_model

## Acknowledgements

This work was funded by NIH grant 1K0ES032458.

