## Supplementary Information for "Addressing spatial misalignment in population health research: a case study of US congressional district political metrics and county health data"

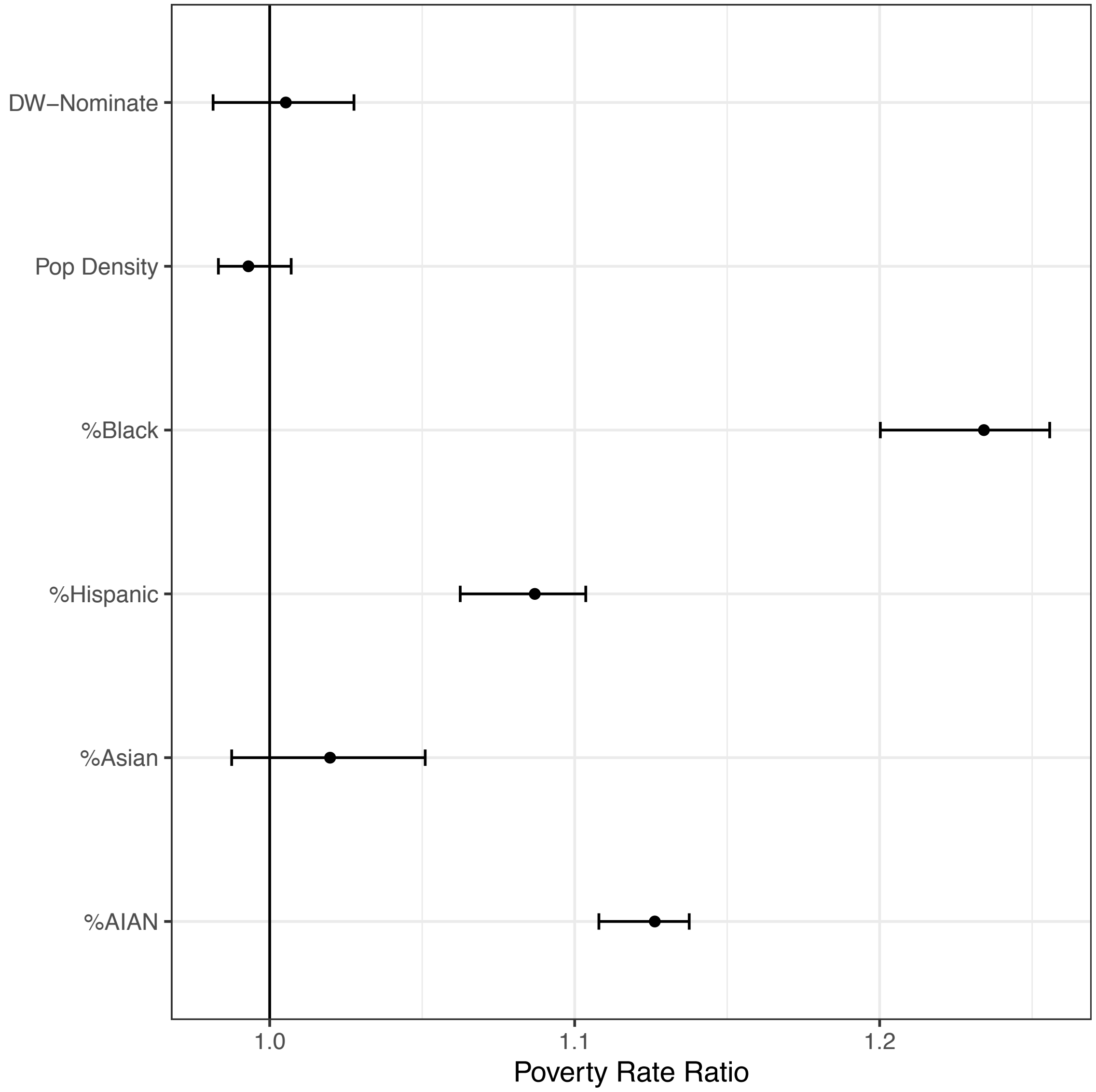


Figure S1. Poverty model rate ratio estimates and 95% credible intervals from the ABRM. All covariates are standardized so that the rate ratio corresponds to a one-standard deviation increase in the coefficients.
